# Adaptation of HL7 FHIR for the exchange of patients’ gene expression profiles

**DOI:** 10.1101/2022.02.11.22270850

**Authors:** Florian Auer, Zhibek Abdykalykova, Dominik Müller, Frank Kramer

## Abstract

High-throughput technologies, especially gene expression analyses can accurately capture the molecular state in patients under different conditions. Thus, their application in clinical routine gains increasing relevance and fosters patient stratification towards individualized treatment decisions. Electronic health records already evolved to capture genomic data within clinical systems and standards like FHIR enable sharing within, and even between institutions. However, FHIR only provides profiles tailored to variations in the molecular sequence, while expression patterns are neglected although being equally important for decision making. Here we provide an exemplary implementation of gene expression profiles of a microarray analysis of patients with acute myeloid leukemia using an adaptation of the FHIR genomics extension. Our results demonstrate how FHIR resources can be facilitated in clinical systems and thereby pave the way for usage for the aggregation and exchange of transcriptomic data in multi-center studies.

## 1. Introduction

Measuring the gene expression in patient samples provides detailed insights into the molecular conditions of the underlying disease. Over the years, high-throughput technologies have evolved to be used in routine clinical diagnostics and foster individualized treatment. At the same time digitalization in healthcare systems advanced to electronic health records (EHR) capturing also genomic data. Interoperability and data sharing between systems and institutions gain more importance with commonly accepted standards like Fast Healthcare Interoperability Resources (FHIR) [1] as a foundation.

FHIR divides the information into modular and extensible components, as well as adapts widely established web standards and the RESTful architecture principle for the sharing of EHRs. Included extensions for genomics data are tailored to cover only variations in the molecular sequence while expression patterns are neglected. Moreover, recommendations for the realization of gene expression results in FHIR are lacking. Nevertheless, these insights are important for decision support and translational research.

Here we provide a feasible FHIR implementation for gene expression profiles from microarray analyses and demonstrate the interoperability of the resulting FHIR resources within an interactive web application.

## 2. Methods

### 2.1. Gene expression data

The data set examines a dose-limiting side effect in patients diagnosed with acute myeloid leukemia (AML) that are treated with chemotherapy [2]. Mucositis, DNA damage within the oral mucosa caused by the chemotherapy is investigated based on the derived gene expression profiles. The samples are collected from punch buccal biopsies from five AML patients pre- and post-chemotherapy, and three healthy controls for comparison. Microarray analysis was performed using Human Genome U133 Plus 2.0 Array (Affymetrix, Santa Clara, CA) with GRCh38.p13 (Genome Reference Consortium Human Build 38, Ensembl release 99) as a reference, followed by a Robust Multichip Average (RMA) normalization of the raw data. The authors made the data available at the EBI Expression Atlas [3] portal by the ID E-GEOD-10746.

We chose this gene expression data set because the conducted analysis represents a typical bioinformatics workflow resulting in several gene expression profiles from the same and different individuals that enable disease classification and patient stratification into risk groups [4].

### 2.2. Adaption in FHIR resources

The central element within FHIR to capture real-world concepts is the *Patient* resource: A study evolves around patient treatment therefore all subsequent patient-specific results, and resources implementing those refer to this base element. Detailed information about the sample donors was not included in the original data set to preserve the anonymity of the participants, instead, we used artificially generated data using Synthea™ [5] to create *Patient* resources as reference. The medical condition of the AML patients was captured by the *Condition* resource to distinguish them from the healthy donors. The single samples are captured by the *Specimen* resource and serve as a link to distinguish between samples collected from the same patient, namely pre- and post-chemotherapy.

The gene expression values are generated based on the GRCh38.p13 reference genome and were measured for each sample. Since all gene expression profiles use the same reference, the single genes contained in the reference genome were included as *MolecularSequence* resources. The actual expression values are treated as single measurements realized as *Observation* resources with the *Observation-geneticsGene* extension referring to the corresponding gene symbol. Although the *Patient* resource is referenced directly within the *Observation* resource, the *Specimen* resource is still required to differentiate between the different samples of the same patient. Using the *MolecularSequence* resource as a reference avoids redundancy of shared genomic information and simplifies the retrieval of the gene expression values for one particular gene across the different samples. An overview of the resources and their links to public databases, as well as references between the resources is shown in figure 1.

**Figure 1.**
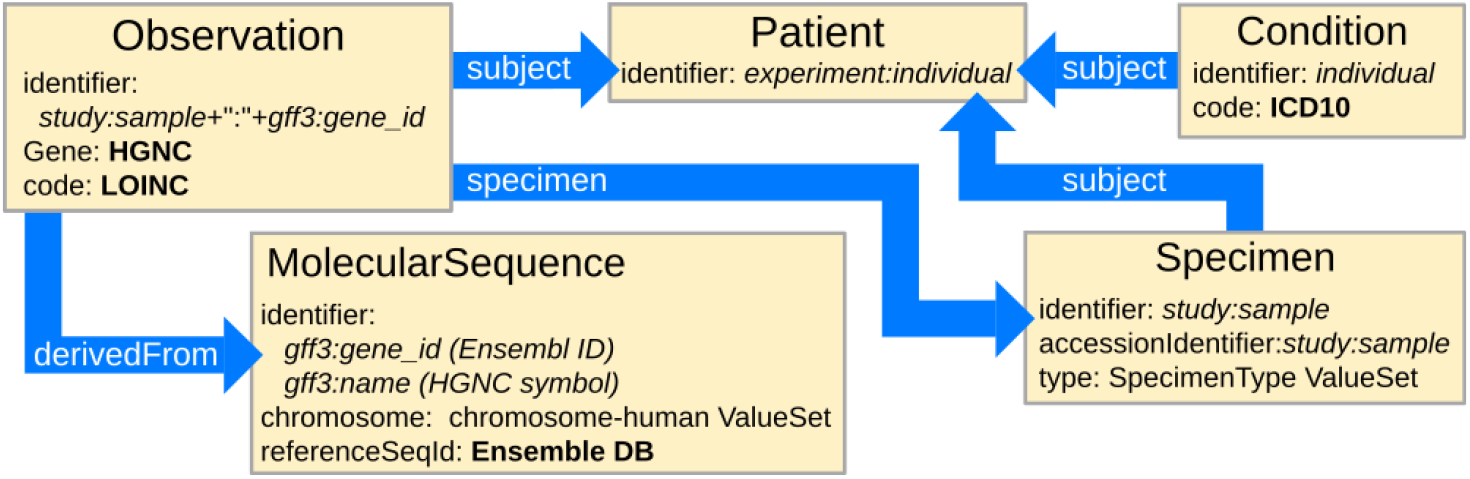
FHIR resources and their references between each other as well as to external databases.

An in-house installation of the dockerized HAPI FHIR server [6] was used for storing the created resources. Additionally, we developed a web application that uses the FHIR REST API to retrieve and display the FHIR resources to demonstrate a minimalistic decision support system.

### 2.3. Data and material availability

All necessary software to reproduce the results is publicly available on GitHub at https://frankkramer-lab/gene-expression-on-fhir. This includes scripts to download the data sets from the official platforms, set up the dockerized HAPI FHIR server and import the data, host the web application, and corresponding source code (GPL-3.0 License). Additionally, a demonstration of the web application with hard-coded excerpts of the FHIR resource data is hosted as a static service using the GitHub pages functionality which can be accessed at https://frankkramer-lab.github.io/gene-expresssion-on-fhir.

## 3. Results

The original data translated to 252,684 resources stored on our FHIR server. For performance improvements, not all gene ids in the reference genome (60,617 ensemble entries) were encoded in FHIR but only those present in the gene expression data. A detailed overview of the created resources and the time requirements is shown in table 1.

**Table 1.** Summary of FHIR resources and time required to upload to the FHIR server.

| FHIR resource | Number of objects | Time for creation |
| --- | --- | --- |
| Patient | 8 | ~1sec |
| Condition | 5 | ~1sec |
| Specimen | 11 | ~2sec |
| MolecularSequence | 21,055 | ~5min |
| Observation | 231,605 | ~45min |

The web application demonstrates the usage of the created resources: Those are obtained directly from the FHIR server, then linked and assembled into a visual representation of the gene expression across the patient samples (figure 2).

**Figure 2.**
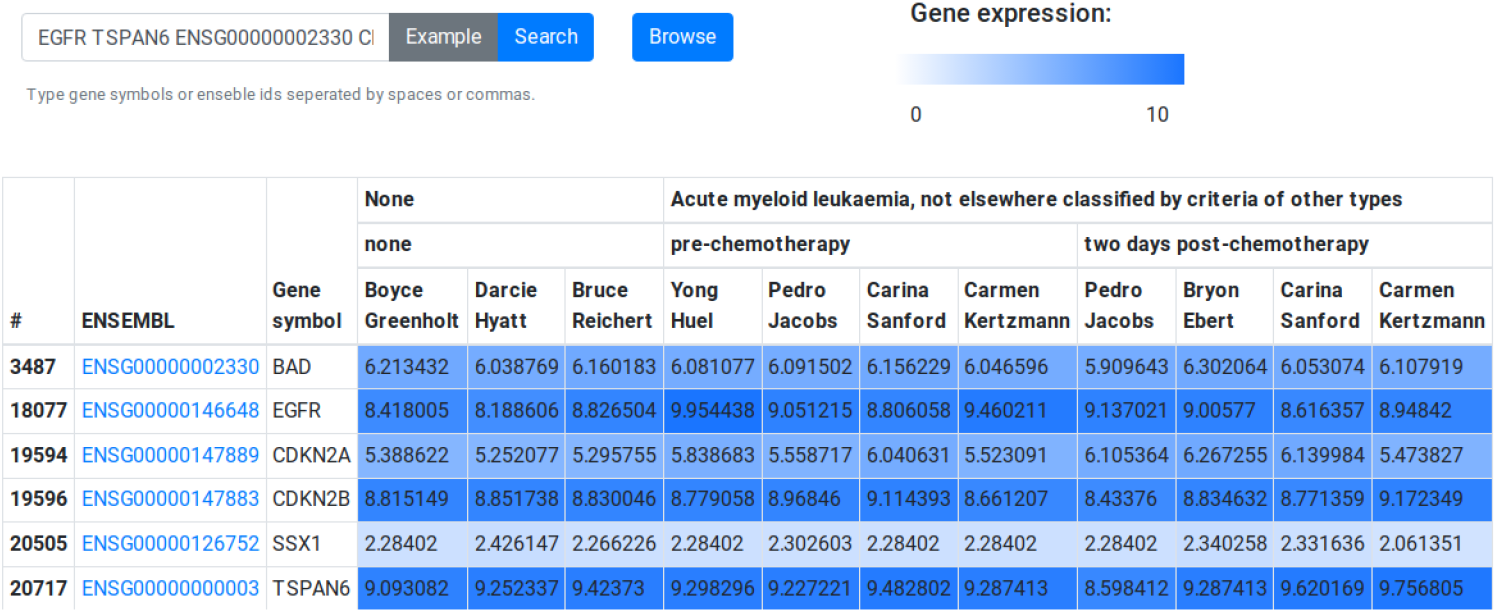
Web application using the created FHIR resources to show the gene expression between the samples of different patients. The heatmap is colored by the corresponding expression intensity.

## 4. Discussion

Through our contribution to the FHIR Genomics extension, we were able to include genomic profiling) data. Since only excerpts of the molecular data are necessary for detailed investigation, FHIR encoded gene expression profiles are suitable for usage in web-based applications. Furthermore, we were able to demonstrate the integration capabilities of FHIR encoded genomic profiles in decision support systems. Further improvements could consist of consolidation of the outcome of the analyses, e.g., significantly differentially expressed genes between samples, as *DiagnosticReport* resources.

## 5. Conclusion

Our results demonstrate how FHIR resources can be facilitated for the clinical exchange of expression profiles. The usage of the adopted resources within our web application demonstrates its feasibility for usage in decision support systems or patient assessment. The further incorporation of genomic features into the FHIR standard offers the opportunity to establish the currently missing standard for the aggregation of various molecular genetics data in a clinical setting. This work contributes to closing this gap and paves the way towards patient stratification through transcriptomic profiling even across health care institutions and within multi-center clinical trials.

## Data Availability

All data produced are available online at https://github.com/frankkramer-lab/gene-expression-on-fhir

https://frankkramer-lab.github.io/gene-expression-on-fhir/

